# Evaluation of a new spike (S) protein based commercial immunoassay for the detection of anti-SARS-CoV-2 IgG

**DOI:** 10.1101/2021.03.10.21253288

**Authors:** Kirsten Alexandra Eberhardt, Felix Dewald, Eva Heger, Lutz Gieselmann, Kanika Vanshylla, Maike Wirtz, Franziska Kleipass, Wibke Johannis, Philipp Schommers, Henning Gruell, Karl August Brensing, Roman-Ulrich Müller, Max Augustin, Clara Lehmann, Manuel Koch, Florian Klein, Veronica Di Cristanziano

## Abstract

**Background:** The investigation of antibody response to SARS-CoV-2 represents a key aspect in facing the COVID-19 pandemic. In the present study, we compared one new and four widely used commercial serological assays for the detection of antibodies targeting S (spike) and NC (nucleocapsid) protein.

**Methods:** Serum samples from a group of apparently non-responders, from an unbiased group of convalescent patients and from a negative control group were sim-ultaneously analyzed by the LIAISON® SARS-CoV-2 S1/S2 IgG test, Euroimmun anti-SARS-CoV-2 S1 IgG ELISA and IDK® anti-SARS-CoV-2 S1 IgG assays. IgG binding NC were detected by the Abbott SARS-CoV-2 IgG assay and by the pan-immunoglobulin immunoassay Elecsys® Anti-SARS-CoV-2. Additionally, samples were also tested by live virus and pseudovirus neutralization tests.

**Results:** Overall, about 50% of convalescent patients with undetectable IgG antibodies using the commercial kit by Euroimmun were identified as IgG positive by Immundiagnostik and Roche. While both assays achieved similarly high sensitivities, Immundiagnostik correlated better with serum neutralizing activity than Roche.

**Conclusions:** Although the proportion of IgG seropositive individuals appears to be higher using more sensitive immunoassays, the protective ability and the potential to serve as indirect markers of other beneficial immune responses warrants for further research.

## Introduction

The investigation of humoral response to SARS-CoV-2 represents a key aspect in facing the COVID-19 pandemic. Although neutralizing antibodies are considered to have an important protective role, the association between seropositivity and immunity, as well as the duration of protective humoral response represent key questions of current research [1-4]. The FDA declared a neutralizing titer ≥ 1:160 as sufficient for plasma donations. However, the definition of an antibody titer conferring protection is still missing [5].

Virus-neutralizing (VN) assays, based on live virus or pseudovirus, are considered the gold standard to conclude on the presence and quantity of specific neutralizing antibodies. However, they are time intensive and require biosafety level 2 or 3 facilities [6]. Most routine diagnostic facilities instead make use of commercially serological immunoassays and high-throughput automated platforms [7]. Various methods for antibody detection are available, including chemiluminescence immunoassay (CLIA), chemiluminescent microparticle immunoassay (CMIA), electrochemiluminescence immunoassay (ECLIA) and enzyme-linked immunosorbent assay (ELISA). They can be divided into assays that recognize specific anti-SARS-CoV-2 antibodies against the spike (S) protein (Figure 1A) with the receptor-binding domain (RBD) and on the other hand in those detecting antibodies targeting the nucleocapsid (NC) protein [8]. Neutralizing antibodies mainly target the RBD domain [6, 9]. Hence, S-protein based assays could be considered more suitable as surrogate for protection [10].

**Figure 1.**
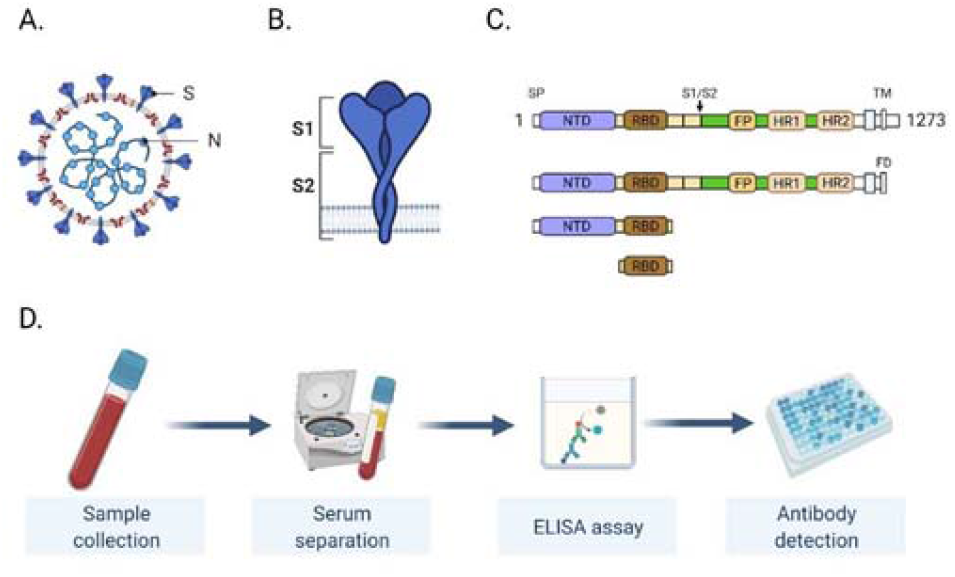
**A)** Schematic drawing of the SARS-CoV-2 virus. Spike (S) and nucleocapsid (N) proteins are highlighted. **B)** Schematic drawing of the trimeric S protein. **C)** The domain structure of the S protein is shown and the three regions which were tested are depicted. All three versions contain the receptor binding domain (RBD). **D)** The ELISA procedure is outlined. SP, signal peptide, NTD, N-terminal domain, S1, spike protein subunit 1, S2, spike protein subunit 2, FP, fusion peptide, HR1, heptad repeat 1 domain, HR2, heptad repeat 2 domain, TM, transmembrane domain, FD, foldon motif

The number of studies providing data about the performance of various serological assays massively increased. So far, the Roche Elecsys, a pan-IG assay for the detection of anti-NC antibodies, was reported to be one of the most sensitive SARS-CoV-2 antibody detection assays [11]. However, it correlated less with neutralizing titers as compared to assays detecting anti-S antibodies [10]. Consequently, despite the ability of the Roche assay to diagnose previous SARS-CoV-2 infection, the potential to predict protection against reinfection is questionable.

A continuous evaluation of commercial assays targeting the S protein has a growing relevance for many reasons. The intensity of antibody response can largely vary in asymptomatic and mild COVID-19 cases, and a relevant proportion of these patients apparently fail to generate a detectable humoral response to SARS-CoV-2 [12-14]. In addition, there are contradictory observations on the persistence of specific antibody levels over time in these groups [15-19]. In case of anti-NC antibodies, a drop in sensitivity over time was already reported for some serological assays [10]. For this reason, it is of outstanding importance to investigate if non detection is equal to absence or just a result of imprecise laboratory assessment methods. Firstly, because seroprevalence could be underestimated [20, 21]. Secondly, considering the results indicating a generation of a stable and potent B memory response, a booster effect regardless of detectable serum antibody levels cannot be excluded as mechanism of protection from infection and/or disease [22, 23]. So far, despite apparently high rates of low- or non-responders, reinfections with SARS-CoV-2 are still reported to be a rare event [24]. Based on these considerations, assay sensitivity could represent a decisive aspect to understand the mechanisms underlying protective humoral responses. Furthermore, highly sensitive S-based protein assays could be relevant for defining time intervals for vaccine boosters, as well as for long term antibody response studies upon vaccination. Finally, the possibility to combine S- and NC-based proteins assays with similar high sensitivity and specificity will be increasingly required to answer the important question of SARS-CoV-2 infection despite active immunization.

Here, we evaluate sensitivity and specificity of a novel S-protein based commercial assay for the detection of serum anti-SARS-CoV-2 IgG. Analyzed samples have been obtained from a cohort of convalescent individuals, mostly recovered from mild COVID-19, recruited at a German University Hospital since April 2020 [16]. In particular, we focused on so-called “non-responders”, here defined as individuals with undetectable serum IgG antibodies as tested with the immunoassay by Euroimmun in our routine laboratory. Additionally, the performance of the new assay by Immundiagnostik was compared to three other IgG qualitative immunoassays, including Roche Elecsys, Abbot (anti-NC IgG), and Diasorin (anti-S1/S2 IgG). Moreover, the correlation between IgG levels and serum neutralizing activity was investigated by live virus and pseudovirus neutralization assays.

## Materials and Methods

### Ethical considerations

All subjects gave their informed consent for inclusion before they participated in the study. The study was conducted in accordance with the Declaration of Helsinki and samples were collected and analyzed under protocols approved by the Institutional Review Board of the University of Cologne, Germany (16-054 and 20-1187).

### Study population

Serum samples were obtained from individuals with molecularly confirmed prior SARS-CoV-2 infection recruited at a German University Hospital between April and September 2020. Most patients recovered from mild COVID-19 and they did not require hospitalization. Two subgroups were investigated: group A included all convalescent patients with previous SARS-CoV-2 infection who attended the clinic between August and September 2020, regardless of their IgG ratio values. Group B was considered as the group of individuals who attended the clinic between April and July 2020 and were tested IgG negative or borderline in our routine screening using the Euroimmun ELISA despite prior SARS-CoV-2 infection. As a last group C, we considered samples collected in 2020 from individuals without any suspicion for SARS-CoV-2 infection, as well as serum samples obtained in 2019 as a negative control group.

### Serological testing for the detection of anti-SARS-CoV-2 antibodies Commercial assays

Serum samples from groups A, B and C were simultaneously analyzed by five different commercial assays for the detection of anti-SARS-CoV-2 antibodies (Supplementary Table 1).

Anti-SARS-CoV-2 IgG targeting the S protein (CLIA) were detected by the LIAISON® SARS-CoV-2 S1/S2 IgG test on the LIAISON® XL (DiaSorin, Vicenza, Italia), by the Euroimmun anti-SARS-CoV-2 IgG ELISA on the Euroimmun Analyzer I (Euroimmun Diagnostik, Lübeck, Germany), and by the IDK® anti-SARS-CoV-2 IgG ELISA (Immundiagnostik AG, Bensheim, Germany) on the DYNEX DSX® (Dynex Technologies, Chantilly, USA). The two latter ELISA assays use the recombinant S1 antigen from of the spike protein (Figure 1).

NC protein antibodies were detected by the SARS-CoV-2 IgG assay (CMIA) provided by Abbott on the Alinity I (Abbott, Abbott Park, Illinois, United States) and by the pan-immunoglobulin immunoassay Elecsys® Anti-SARS-CoV-2 (ECLIA) on the Cobas e (Roche Diagnostics, Mannheim, Germany).

All assays were interpreted according to manufacturer´s recommendations. IgG values by Immundiagnostik were capped at 3.8 OD at the upper end for analysis. In case of the assays by Euroimmun and DiaSorin, borderline results were counted as negative. However, we alternatively recalculated our findings counting borderline results as positive to evaluate if test performances would profoundly differ.

### Live virus assay to determine SARS-CoV-2 neutralizing activity (LVN)

After inactivation of complement by heating at 56° for 30 minutes, serum samples were diluted 1:10 and 1:50 in 2-folds in DMEM medium (Dulbecco Dulbecco’s Modified Eagle’s Medium, Gibco, Dublin, Ireland) and mixed with 100 TCID50 (Tissue culture infectious dose 50) of wild type virus (isolated from a patient at the University Hospital of Cologne) to a volume of 100 µL. Virus-serum mixture was incubated for one hour at 37°C. Afterwards, 50 μl of VERO E6 cell suspension (250.000 cells/ml) were added to each sample dilution. Cell plates were incubated at 37° for 4 days before to microscopically determine the virus-related cytopathic effects (CPE).

### Pseudovirus assay to determine SARS-CoV-2 neutralizing activity (PVN)

For testing SARS-CoV-2 neutralizing activity using pseudovirus, serial dilutions of serum (heat inactivated at 56°C for 45 min) were coincubated with pseudovirus supernatants for 1 h at 37°C and thereafter, 293T cells engineered to express ACE2 were added [25]. After a 48-hour incubation at 37°C and 5% CO2, luciferase activity was determined after addition of luciferin/lysis buffer (10 mM MgCl2, 0.3 mM ATP, 0.5 mM Coenzyme A, 17 mM IGE-PAL (all Sigma-Aldrich), and 1 mM D-Luciferin (GoldBio) in Tris-HCL) using a micro-plate reader (Berthold). After subtracting background relative luminescence units (RLUs) of un-infected cells, 50% Inhibitory dose (ID50) was determined as the serum dilution with 50% RLU reduction compared to untreated virus control wells. Every serum sample was measured on different days in two independent experiments and the mean ID50 values is presented.

### Statistical Analysis

Continuous variables were expressed as median (interquartile range, IQR) or mean ± standard deviation (SD) and compared using the Wilcoxon rank sum test or the unpaired Student’s t-test. Categorical variables were compared using either the χ2 test or the Fisher exact test, as appropriate. The Spearman rank correlation coefficient ρ was calculated as measure of strength of the relationship between serological assay outcomes and the pseudovirus neutralization assay. The correlation between the 3-level ordinal live virus neutralization assay outcome and categorized binary outcomes of serologic assays, was evaluated by calculating the Kendall’s coefficient of rank correlation τ. Additionally, two neutralizing cutoff titers were assessed for their concordance with the binary outcomes of the commercial serological assays using the Cohen’s κ. Two-sided p-values were presented, and an α of 0.05 was determined as the cutoff for significance. All statistical analyses were performed using R (version 3.6.3, R Foundation for Statistical Computing, Vienna, Austria).

## Results

### Establishment of a new SARS-CoV-2 serological test

Serological tests mainly focus on the S and NC protein from the SARS-CoV-2 virus (Figure 1A, 1B). As the S protein might be the most crucial protein for the infection, different regions of the S protein have been compared and evaluated for the use in an ELISA kit. Three different regions, S protein ectodomain, RBD, and S1 truncated, have been recombinantly expressed in HEK293 cells and intensively tested (Figure 1C). The S1 truncated was chosen for further usage since it performed best in different tests. According to Immundiagnostik, 7 out of 762 plasma samples, collected between 2017 and 2018, tested positive for the SARS-CoV-2 by ELISA. Furthermore, no cross reactivity to plasma probes for Adenovirus, Epstein Barr-Virus, Influenza A/B, HCoV-229E, HCoV-HKU1, HCoV-NL63, and HCoV-OC43 was detected. Additional information can be obtained from in the Immundiagnostik manual.

### Sensitivity of five commercially available serological tests and two virus-neutralizing immunoassays in a cohort with previous infection with SARS-CoV-2 (group A)

To evaluate the general sensitivity of serological tests with different antigen targets, we compared three commercially available immunoassays targeting the S protein, two assays detecting the NC protein, the combination of assays targeting different antigens, and two VN assays (Supplementary Table 2). Serum samples of 363 convalescent patients with prior SARS-CoV-2 infection collected between August and September 2020 (group A) were used for this evaluation (Figure 2A). In this group 5.9% of the individuals declared themselves as asymptomatic, 89.8% participants had a mild course of disease, while 4.33% of the subjects were hospitalized because of COVID-19. The median time between infection and antibody determination was 154 days (Table 1). As indicated by Figure 2B, the sensitivities achieved by immunoassays targeting the S protein ranged between 77.1% and 89.2% with the assay by Immundiagnostik being the most sensitive in that group. The serological tests detecting the NC antigen performed vastly different. While the test by Roche achieved the highest sensitivity (93.1%) of all assays, the Abbott immunoassay only reached 53.7% sensitivity in this general cohort of recovered individuals. A combination of the best performing immunoassays with different protein targets resulted in a sensitivity of 93.9%. Interestingly, the sensitivity of 91% achieved by the VN assay was below the sensitivity of the commercially available Roche test but higher than all the S protein targeting tests. 342 (94.2%) participants with past SARS-CoV-2 infection were tested IgG positive in at least one of the immunoassays evaluated.

**Figure 2.**
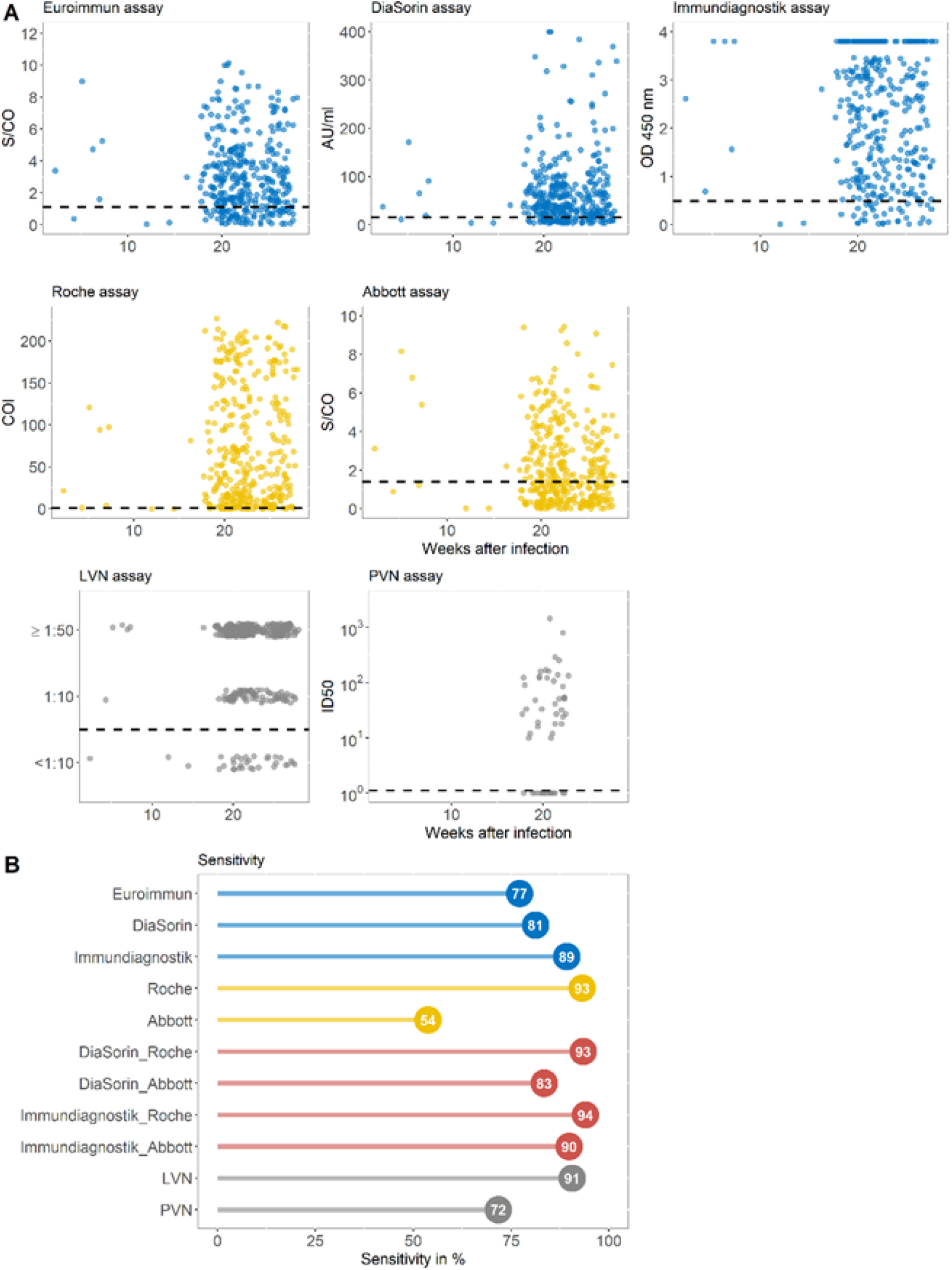
**A)** IgG ratio values of five commercially available immunoassays targeting the S (spike) (blue) or the NC (nucleocapsid) protein (yellow) and virus-neutralizing assays (grey) displayed against the weeks after infection for each individual from group A. Dashed horizontal lines display cutoff values of individual serological assays. Results of the virus-neutralizing immunoassay are categorized into <1:10, 1:10, and >=1:50. **B)** The sensitivity in % achieved by assays against the S protein (blue), the NC antigen (yellow), the combination of assays targeting the different proteins (red), and the VN test (grey) for group LVN, live virus neutralization assay; PVN, pseudovirus neutralization assay

**Table 1.** Demographical and clinical characteristics of participants.

| Parameters | Group A ,<br>n=363 | Group B ,<br>n=169 |
| --- | --- | --- |
| Female, n (%) | 200 (55.24) | 103 (61.68) |
| Male, n (%)* | 162 (44.75) | 64 (38.32) |
| Age in years, mean ± SD | 44.09 ± 12.86 | 42.69 ± 12.86 |
| Days after disease onset, median (IQR) | 154 (141-176) | 47 (35-56) |
| Asymptomatic, n (%) | 15 (5.90) | 20 (16.00) |
| Mild, n (%) | 228 (89.77) | 100 (80.00) |
| Severe, n (%)** | 11 (4.33) | 5 (4.00) |

*SD, standard deviation; IQR, interquartile range; \* no data available for 1 subject in group A and 2 subjects in group B; \*\* no data available for 109 subjects in group A and 44 subjects in group B*

Figure 3A displays the correlation between the different commercial immunoassays and the pseudovirus neutralization assay. The relationship between assays targeting the S domain is strong with a Spearman rank correlation coefficient ρ ranging between 0.80 and 0.85. In contrast to this, the relationship between the assays targeting the NC protein is slightly less strong with rho=0.58-0.65. The relation between live virus neutralization assay results and those from the different commercial immunoassays are shown in Figure 3 B. The Kendall’s τ between the LVN assay and the serological assay ranges between 0.40 and 0.65 with higher values for the assays targeting S protein antigens. Cohen’s κ as a measure of concordance between neutralizing titer cutoffs and binary results of serological assays ranged widely and reached the highest value of 0.71 in case of the Immundiagnostik assay at a LVN-cutoff titer of 1:10 and of the Euroimmun assay at a LVN cut-off of 1:50 (κ=0.61).

**Figure 3.**
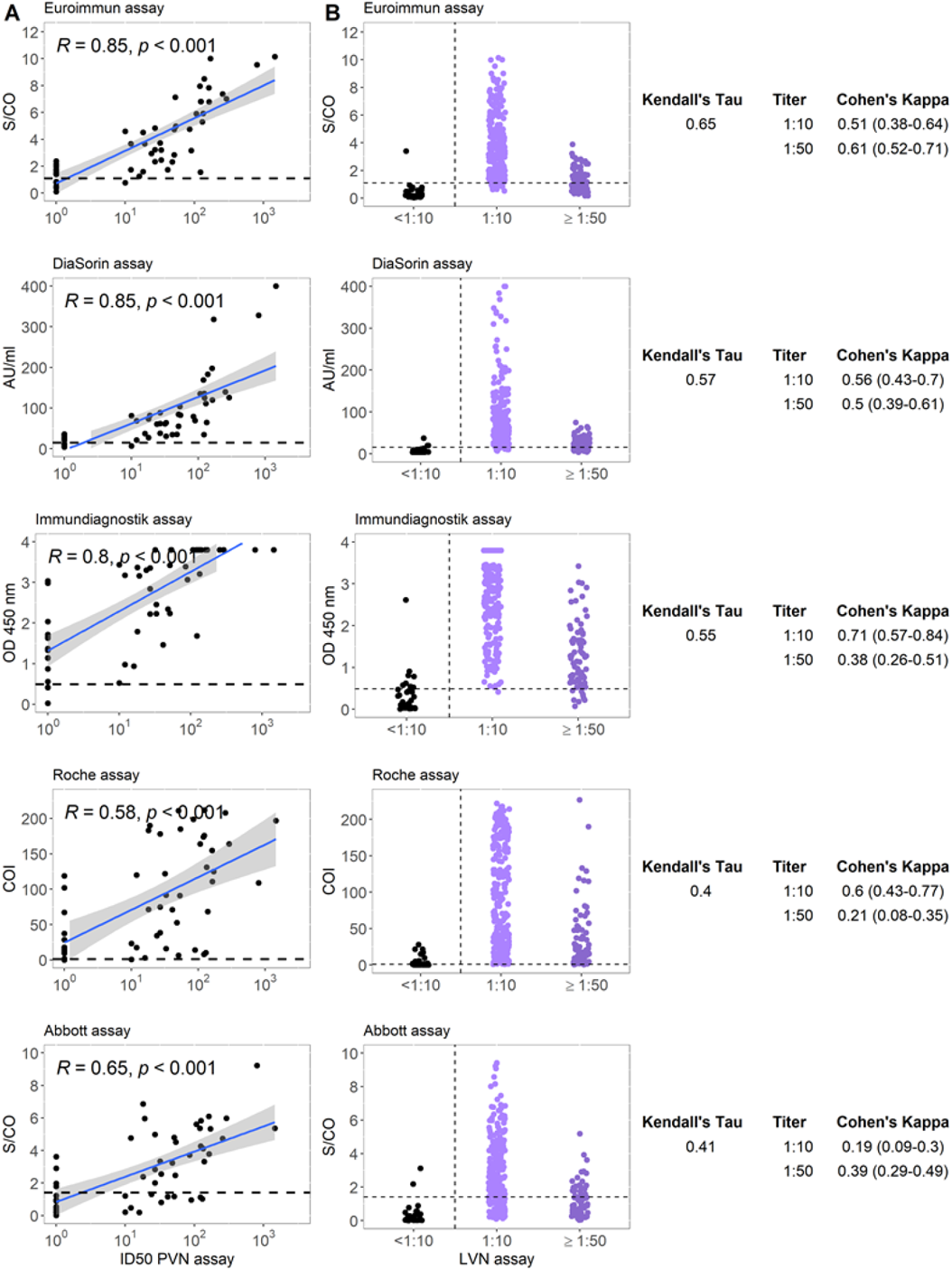
**A)** Correlation between five commercial anti-SARS-CoV-2 serological assays and pseudo-virus-neutralizing antibody titer for group A. R represents the Spearman rank correlation coefficient ρ. **B)** Correlation between five commercial anti-SARS-CoV-2 serological assays and live virus neutralizing antibody titer for group A. Horizontal lines represent cutoff values for individual commercial tests. Kendall’s τ and Cohen’s κ are displayed for each test combination. LVN, live virus neutralization assay; PVN, pseudovirus neutralization assay Sensitivity of four commercially available serological tests and two virus-neutralizing immunoassays in a cohort of non-responders (group B)

Between April and July 2020, 169 individuals with prior SARS-CoV-2 infection attending our clinic were classified as non-responders in the routine IgG screening using the Euroimmun ELISA (group B). In this group 16.0% of the subject were asymptomatic, 80.0% participants had a mild course of disease, and 4.0% of the subjects were hospitalized because of COVID-19. The median time interval between infection and antibody determination was 47 days (Table 1).

By retesting the samples with two other assays against the S protein, two commercially available immunoassays targeting the NC protein, different test combinations, and two VN assays, we investigated which serologic test would perform best in detecting IgG seropositive individuals in this apparently non-responder population (Figure 4A and Supplementary Table 3).

**Figure 4.**
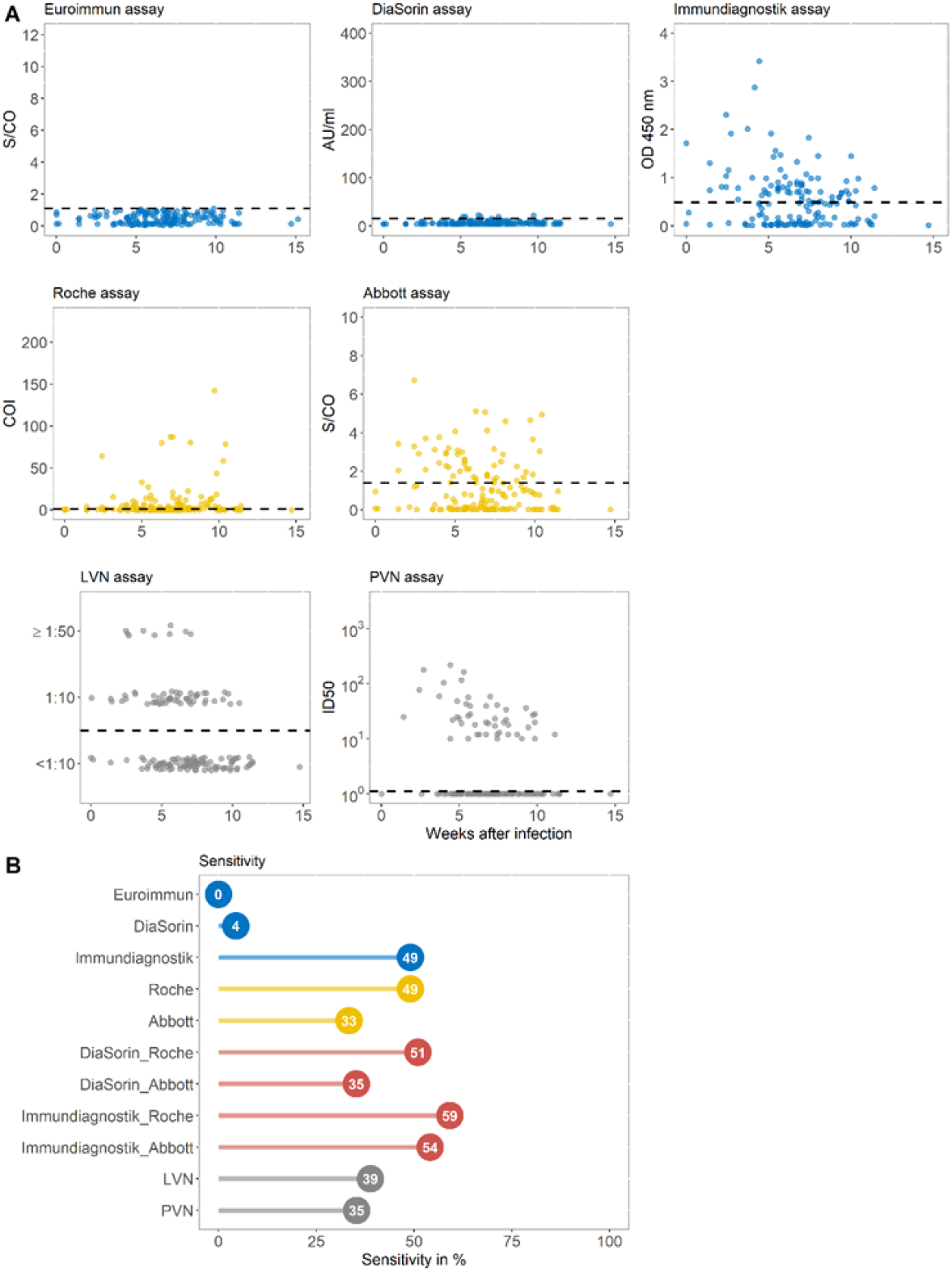
**A)** IgG ratio values of five commercially available immunoassays targeting the S (spike) (blue) or the NC (nucleocapsid) protein (yellow) and virus-neutralizing assays (grey) displayed against the weeks after infection for each individual from group B. Dashed horizontal lines display cutoff values of individual serological assays. Results of the virus-neutralizing immunoassay are categorized into <1:10, 1:10, and >=1:50. **B)** The sensitivity in % achieved by assays against the S protein (blue), the NC antigen (yellow), the combination of assays targeting the different proteins (red), and the VN test (grey) for group LVN, live virus neutralization assay; PVN, pseudovirus neutralization assay

While the DiaSorin immunoassay only detected very few patients as IgG positive (4.4%), the test by Immundiagnostik found almost the half (49.06%) of this non-responder cohort to be IgG positive. Exactly the same number of apparently non-responders (49.06%) were found to be IgG positive using the Roche test. Remarkably, when combining the most sensitive tests with different target proteins (Immundiagnostik and Roche), 94 out of 159 patients (59.1%) were detected as seroconverted instead of being IgG negative as assumed after routine screening with the Euroimmun assay (Figure 4B). Using two different virus-neutralizing immunoassays, 38.8% and 35.3% of initially IgG negative participants were tested positive for the presence of neutralizing antibodies. 97 (61%) individuals with past SARS-CoV-2 infection but undetectable IgG antibodies by the Euroimmun ELISA were classified as seropositive in at least one of the other assays evaluated.

The correlation between the different commercial immunoassays and the pseudovirus neutralization assay for the special sub-cohort of apparently non-responders is displayed in Figure 5A. The relationships between the PVN assay and all evaluated commercial assays is moderate with a Spearman rank correlation coefficient ρ ranging between 0.53 in case of the Roche assay and 0.66 for the assay by Immundiagnostik. The relation between neutralization assay using live virus results and those from the different commercial immunoassays in the cohort of Euroimmun non-responders are shown in Figure 5B. The Kendall’s τ between the ordinal VN assay and the categorized serological tests ranges largely between 0.12 and 0.66 with the highest correlation observed with the Immundiagnostik assay. The correlation coefficients between the LVN assay and the serological assays targeting the NC protein do not differ profoundly from those measured in the unbiased convalescent cohort. The highest concordance as determined by the Cohen’s κ was observed at an LVN-titer of 1:10 with the serologic assay of Immundiagnostik (κ=0.67), whereas it was negative (κ=-0.05) in case of the DiaSorin assay at a neutralizing antibody cutoff-titer of 1:50 (Figure 5B).

**Figure 5.**
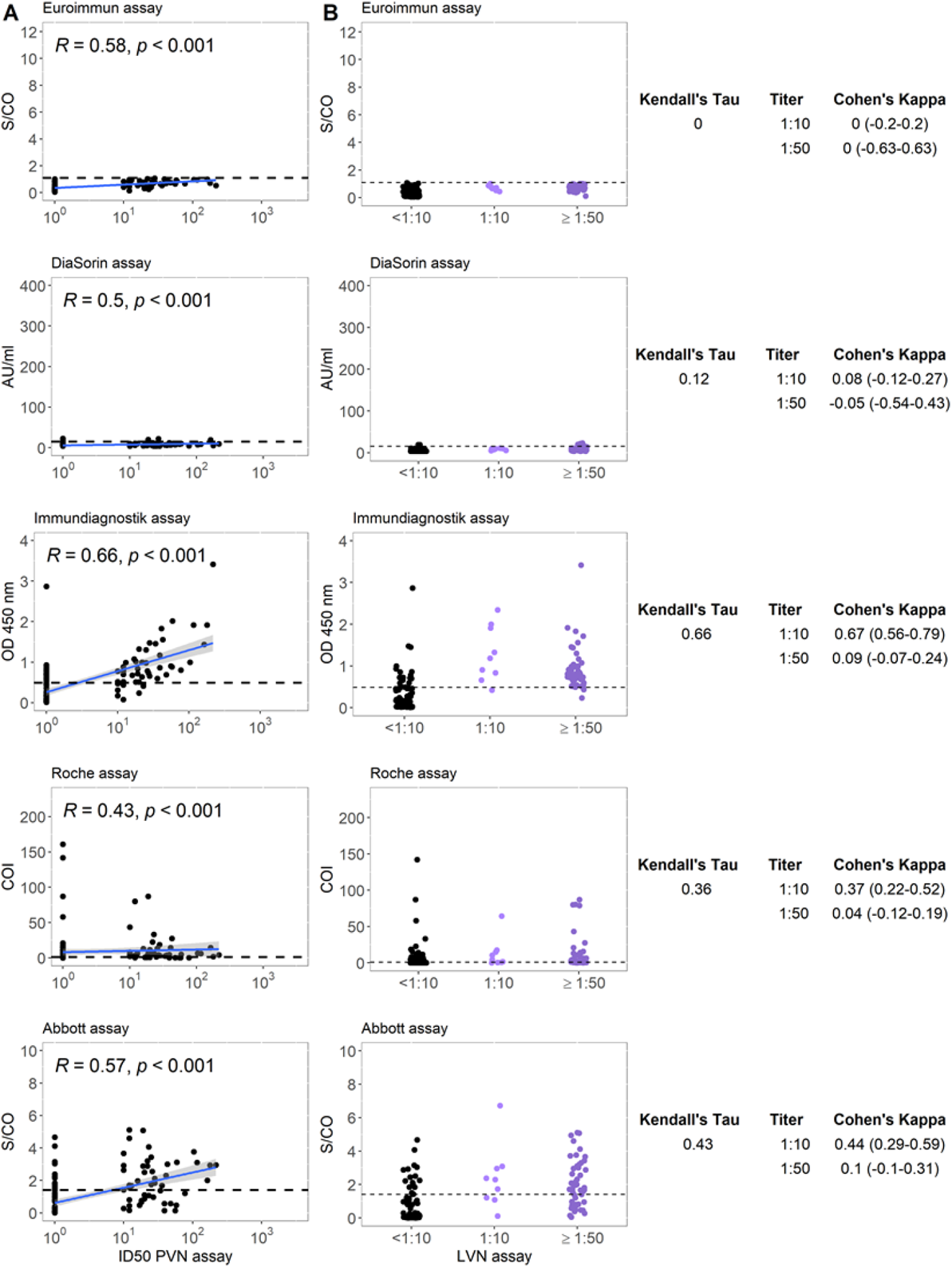
**A)** Correlation between five commercial anti-SARS-CoV-2 serological assays and pseudo-virus-neutralizing antibody titer for group B. R represents the Spearman rank correlation coefficient ρ. **B)** Correlation between five commercial anti-SARS-CoV-2 serological assays and live virus neutralizing antibody titer for group B. Horizontal lines represent cutoff values for individual commercial tests. Kendall’s τ and Cohen’s κ are displayed for each test combination. LVN, live virus neutralization assay; PVN, pseudovirus neutralization assay Specificity of five commercially available serological tests and one virus-neutralizing immunoassay in a SARS-CoV-2 negative control group (group C)

We compared the performance in terms of specificity of five serological tests with different antigen targets and one virus-neutralizing assay using a control group (group C, n=227) with 177 serum samples of individuals without suspected SARS-CoV-2 infection collected in 2020 and 50 serum samples that were collected in 2019 in our clinic. All assays or combination of those achieved a specificity of 99-100% (Figure 6 and Supplementary Table 4).

**Figure 6.**
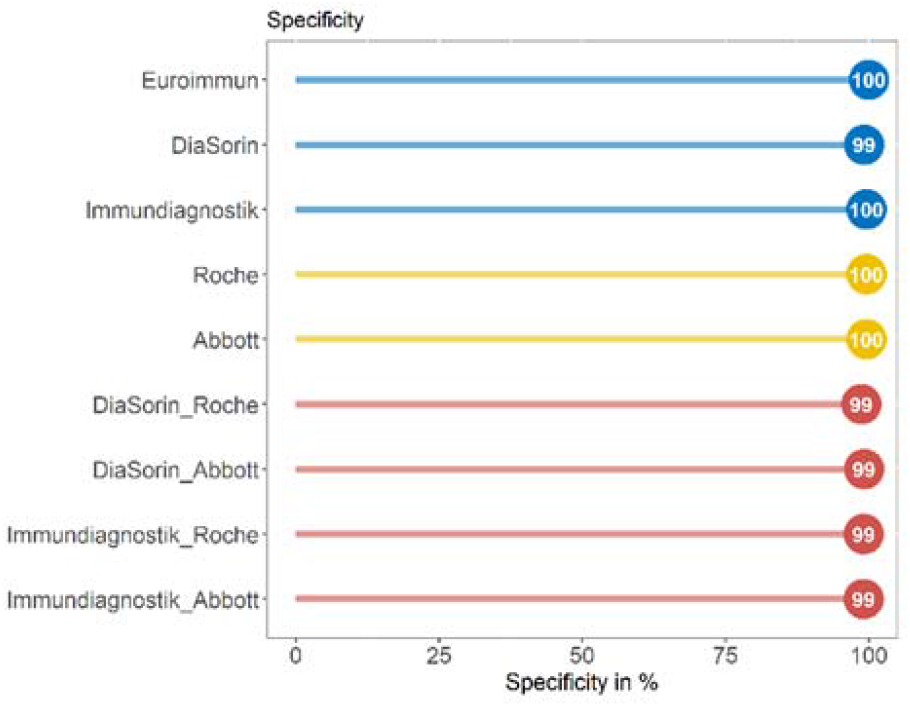
The specify achieved by assays against the S (spike) protein (blue), the NC (nucleocapsid) antigen (yellow), and the combination of assays targeting the different proteins (red) for group C.

## Discussion

The detection of specific antibodies against defined infectious pathogens is commonly used as marker of infection or protection. The determination of the serological immune status against hepatitis B virus or measles are widely known examples of tests to evaluate individual protection against these agents [26]. However, in case of SARS-CoV-2, it was observed that profound proportions of patients can remain seronegative even moths after infection or that antibody levels wane over time, particularly in asymptomatic or mild courses of disease [27-31]. Considering that the majority of COVID-19 cases have a mild symptomatic course of disease, it is of outstanding importance to well characterize individuals with low or undetectable serum IgG response and investigate, if non-detection is equal to absence or just a result of imprecise laboratory assessment methods [12].

In the present study, we evaluated the performance of a novel ELISA commercial assay approved for the detection of anti-SARS-CoV-2 IgG targeting epitopes within the S1 region in patients recovered from mostly mild COVID-19. In particular, we studied two different sub-cohorts of convalescent individuals, one unbiased group of recovered patients and one group of apparently serological non-responders that were tested IgG seronegative by Euroimmun.

To our knowledge, this is the first study investigating the performance of common serological commercial assays in a target non-responder group recovered from mild COVID-19.

The evaluation of serologic immunoassays in the general cohort of individuals with previous SARS-CoV-2 infection unraveled varying test performances. In line with other works, tests targeting the S protein ranged between a sensitivity of 77.1% in case of the Euroimmun IgG ELISA [32], and 89.2% achieved by the new serologic test by Immundiagnostik. Differently from previous data, the Abbott Alinity IgG assay achieved a profoundly lower sensitivity compared to the Roche immunoassay (53.7% and 93.1%, respectively) [33].

In the sub-cohort of apparently non-responders, defined as individuals with undetectable IgG antibodies by the Euroimmun IgG ELISA despite confirmed prior SARS-CoV-2 infection, we detected a remarkably high proportion of patients that produced specific an-ti-SARS-CoV-2 IgG antibodies. Whereas the assay by DiaSorin detected IgG in only 4% of these patients, the Abbot assay was able to detect antibodies in 33% of the same individuals. The tests by Immundiagnostik and Roche, both found 49.1% of the apparently non-responders to produce IgG serum antibodies against SARS-CoV-2. When combining these two immunoassays with the highest detection rate, this proportion reached even 59.1%. This noticeable result confirms that almost 60% of the participants tested seronegative by Euroimmun are instead IgG seropositive.

The moderate performance of the Allinity Abbot assay in our study is in line with the results by Muecksch et al. [10]. The authors reported a decay of the sensitivity from >90% within the first 40 days post-infection to 71% in samples tested more than 80 days after diagnosis when tested with the Abbott immunoassay, whereas Roche Elecsys titers stayed stable overtime. Similarly, in the present study, the Abbot assay was shown to be much less sensitive in comparison to all other commercial assays in the unbiased cohort of convalescent patients (group A) who were tested after a median time of 154 days post-infection. In contrast to this, the difference between the Abbott and the Roche assays was less prominent in the group of apparently “non-responders” (group B). This group was tested for the presence of antibodies already after a median time of 47 days after SARS-CoV-2 infection. Muecksch et al. suggested that the different performance of the two NC assays may be attributable to the antigen bridging approach characterizing the Roche assay, which measures the evolution of antibody binding properties despite their decay in total amount over time. Therefore, it is advisable to prefer the technology by Roche for the detection of NC IgG antibodies.

Overall, these data confirmed previous conclusions that disease severity and time since infection can critically impact on the declared sensitivity of commercial SARS-CoV-2 serological assays. Furthermore, the S protein appears to be a more reliable target than the NC region [20, 34].

Our data suggest that the performance of the Immundiagnostik assay in terms of sensitivity is similarly high compared to the assay by Roche in both subgroups. This indicates that lower antibody concentrations do not imply a loss of sensitivity despite the high specificity of both assays. However, the Immundiagnostik assays might be preferred over the one by Roche for the practical reasons that it can be used manually or by different automated platforms without the issue of a short onboard stability as it is the case for the Roche kit. Among the S-protein based assays, Immundiagnostik achieved the highest sensitivity. A potential explanation for the superiority of the new serologic test by Immundiagnostik compared to the widely used IgG Euroimmun, both targeting S1, could be that different parts of the S-protein from SARS CoV-2 were used. For the development of the Immundiagnostik ELISA kit, three different regions of the S-protein were tested. The most robust signal with low background noise and high reproducibility was obtained with the N-terminal part of the S1-protein. For the set-up, over 762 plasma samples, collected between 2017 and 2018, were tested to define a tight cut off. In comparison to the Euroimmun S1 protein, the Immundiagnostik protein is 156 amino acid shorter and the common region differs by four amino acids.

Overall, neutralizing antibodies are considered one of the main parameters to measure protective immunity against SARS-CoV-2, although their role in case of ongoing disease is discussed more controversial [35-38]. Thus, the detection of neutralizing antibodies could play a key role in monitoring vaccine efficacy but since biosafety requirements are high, it is of outstanding interest that new serologic immunoassays are not only more sensitive, but also correlate well with neutralization assays [39-41]. Recent studies indicated that commercial assays targeting the S protein often correlate better with neutralizing antibody titers than those targeting the NC protein [42, 43]. Likewise, our presented data suggest that in general populations of individuals recovered from SARS-CoV-2 infection, immunoassays targeting the S antigen correlated to a higher degree with results from the virus neutralizing assay than those targeting the NC protein. In the special group of “non-responders”, the serologic test by Immundiagnostik achieved the highest correlation with results from the virus neutralizing assay, and the concordance was highest if low neutralizing titers of 1:10 were considered as cutoff value for positivity. On the other hand, the Euroimmun assay achieved a better concordance if a cutoff of 1:50 for neutralization was chosen in the unbiased group. These results confirm the higher sensitivity of Immundiagnostik assay and the ability to detect low-level neutralizing antibodies but it is unclear which serum antibodies titers can be considered as protective. Although the proportion of IgG seropositive individuals appears to be higher if tested with more sensitive immunoassays, which are at the same time highly specific, the potential of these low antibody levels to serve as markers of beneficial immune functions remains to be further investigated. Recent studies found that the memory B cell compartment remains stable with the ability to generate potent neutralizing antibodies upon re-exposure to SARS-CoV-2 and thus might represent a more suitable marker for humoral immune response than antibody titers [23, 44].

## Conclusions

Our study has two main outcomes. Firstly, we found the serological assays by Immundiagnostik and Roche to be the most sensitive commercial tests from those evaluated. Their combined use would be for example an advisable option in order to differentiate infected from vaccinated. Secondly, we found that more than half of those participants who recovered from SARS-CoV-2 infection but with undetectable IgG antibodies using the commercial test by Euroimmun to be IgG seropositive when tested by Roche and Immundiagnostik. Compared to Roche, the new serological assay by Immundiagnostik correlated better with serum neutralizing activity, particularly in patients with only low antibody concentrations after presumably mild infection. The protective role of detectable low-level neutralizing activity deserves further investigation, considering the increasing data pointing to the role of memory B cell compartment.

## Data Availability

Dataset analyzed during the current study is available in supplementary materials.

## Supplementary Materials

Supplementary Table 1: Dataset; Supplementary Table 2: Sensitivity of five commercially available serological tests, combination of tests, and one virus-neutralizing immunoassay in a cohort with previous infection with SARS-CoV-2.; Supplementary Table 3: Sensitivity of four commercially available serological tests, combination of tests, and one virus-neutralizing immunoassay in a cohort with previous infection with SARS-CoV-2 but undetectable IgG antibodies by the Euroimmun assay; Supplementary Table 4: Specificity of five commercially available serological tests, combination of tests, and one virus-neutralizing immunoassay in a SARS-CoV-2 negative control group.

## Author Contributions

Conceptualization, Kirsten Alexandra Eberhardt, Felix Dewald, Kanika Vanshylla, Manuel Koch, Florian Klein and Veronica Di Cristanziano; Formal analysis, Kirsten Alexandra Eberhardt, Felix Dewald and Veronica Di Cristanziano; Investigation, Kirsten Alexandra Eberhardt, Felix Dewald, Eva Heger, Lutz Gieselmann, Kanika Vanshylla, Maike Wirtz, Franziska Kleipass, Philipp Schommers, Henning Gruell, Karl August Brensing, Roman-Ulrich Müller, Max Augustin, Clara Lehmann and Veronica Di Cristanziano; Methodology, Felix Dewald, Manuel Koch and Veronica Di Cristanziano; Project administration, Manuel Koch and Florian Klein; Resources, Wibke Johannis, Manuel Koch and Florian Klein; Supervision, Florian Klein and Veronica Di Cristanziano; Writing – original draft, Kirsten Alexandra Eberhardt, Felix Dewald and Veronica Di Cristanziano; Writing – review & editing, Eva Heger, Lutz Gieselmann, Kanika Vanshylla, Maike Wirtz, Franziska Kleipass, Wibke Johannis, Philipp Schommers, Henning Gruell, Karl August Brensing, Roman-Ulrich Müller, Max Augustin, Clara Lehmann, Manuel Koch and Florian Klein.

## Funding

This work was funded by grants from the German Center for Infection Research (DZIF to F.Klein), the German Research Foundation (DFG; CRC 1279, F.Klein; CRC 1310, F.Klein; FOR2722, M.K.), the European Research Council (ERC-StG639961, F.Klein), and COVIM: „NaFoUniMedCovid19” (FKZ: 01KX2021).

## Data Availability Statement

All observation data are accessible via the Supplementary Materials.

## Acknowledgments

We thank all participants contributing to this study. Furthermore,we owe special thanks to Dr. Franz Paul Armbruster, Claudia Schumann, Sabine Friedl, Dagmar Szellas, Guerino Rizzo, and Dr. Eva-Maria Rogg from Immundiagnostik for establishing the ELISA kit in collaboration with the University Hospital Cologne. Figure 1 was created with BioRender.com.

## Conflicts of Interest

The University of Cologne (Medical Faculty) receives royalties for the co-development of the IDK® anti-SARS-CoV-2 IgG ELISA and IDK® anti-SARS-CoV-2 IgM ELISA kits. IDK® had no role in the design of the study; in the collection, analyses, or interpretation of data; in the writing of the manuscript, or in the decision to publish the results.

## Notes

### Competing Interest Statement

The University of Cologne (Medical Faculty) receives royalties from Immundiagnostik (IDK) for the co-development of the IDK anti-SARS-CoV-2 IgG ELISA and IDK anti-SARS-CoV-2 IgM ELISA kits. However, until now no royalty has been paid. We expect within the next months some royalties being paid.
IDK had no role in the design of the study; in the collection, analyses, or interpretation of data; in the writing of the manuscript, or in the decision to publish the results.

### Funding Statement

This work was funded by grants from the German Center for Infection Research (DZIF to F.K.), the German Research Foundation (DFG; CRC 1279, F.K.; CRC 1310, F.K.; FOR2722, M.K.), the European Research Council (ERC-StG639961, F.K.), and COVIM: NaFoUniMedCovid19 (FKZ: 01KX2021).

### Author Declarations

Samples were collected and analyzed under protocols approved by the Institutional Review Board of the University of Cologne, Germany (16-054 and 20-1187)

