## Supplementary Table 1; Supplementary Table 2; Supplementary Table 3; Supplementary Table 4 for "Evaluation of a new spike (S) protein based commercial immunoassay for the detection of anti-SARS-CoV-2 IgG": Supplementary Materials.docx

**Supplementary Table 2.** Sensitivity of five commercially available serological tests, combination of tests, and one virus-neutralizing immunoassay in a cohort with previous infection with SARS-CoV-2.

|  | n | Pos/neg | Sensitivity in % |
| --- | --- | --- | --- |
| Euroimmun | 363 | 280/83 | 77.13 |
| DiaSorin | 363 | 295/68 | 81.27 |
| Immundiagnostik | 362 | 323/39 | 89.23 |
| Roche | 363 | 338/25 | 93.11 |
| Abbott | 363 | 195/168 | 53.71 |
| Euroimmun/Roche | 363 | 338/25 | 93.11 |
| Euroimmun/ Abbott | 363 | 292/71 | 80.44 |
| DiaSorin /Roche | 363 | 339/24 | 93.39 |
| DiaSorin / Abbott | 363 | 303/60 | 83.47 |
| Immundiagnostik/Roche | 363 | 341/22 | 93.94 |
| Immundiagnostik/ Abbott | 363 | 326/37 | 89.81 |
| Live virus neutralization test | 351 | 318/33 | 90.60 |
| Pseudovirus neutralization test | 53 | 38/15 | 71.70 |

**Supplementary Table 3.** Sensitivity of four commercially available serological tests, combination of tests, and one virus-neutralizing immunoassay in a cohort with previous infection with SARS-CoV-2 but undetectable IgG antibodies by the Euroimmun assay.

|  | n | Pos/neg | Sensitivity in % |
| --- | --- | --- | --- |
| Euroimmun | 159 | 0/159 | 0 |
| DiaSorin | 159 | 7/152 | 4.40 |
| Immundiagnostik | 159 | 78/81 | 49.06 |
| Roche | 159 | 78/81 | 49.06 |
| Abbott | 159 | 53/106 | 33.33 |
| DiaSorin/Roche | 159 | 81/78 | 50.94 |
| DiaSorin/Abbott | 159 | 56/103 | 35.22 |
| Immundiagnostik/Roche | 159 | 94/65 | 59.12 |
| Immundiagnostik/Abbott | 159 | 86/73 | 54.09 |
| Live virus neutralization test | 152 | 60/93 | 38.82 |
| Pseudovirus neutralization test | 136 | 88/48 | 35.29 |

**Supplementary Table 4.** Specificity of five commercially available serological tests, combination of tests, and one virus-neutralizing immunoassay in a SARS-CoV-2 negative control group.

|  | n | Pos/neg | Specificity in % |
| --- | --- | --- | --- |
| Euroimmun | 227 | 0/227 | 100.0 |
| DiaSorin | 227 | 2/225 | 99.1 |
| Immundiagnostik | 226 | 1/225 | 99.6 |
| Roche | 227 | 1/226 | 99.6 |
| Abbott | 227 | 1/226 | 99.6 |
| Euroimmun/Roche | 227 | 1/226 | 99.6 |
| Euroimmun/Abbott | 227 | 1/226 | 99.6 |
| DiaSorin /Roche | 227 | 3/224 | 98.7 |
| DiaSorin/Abbott | 227 | 2/225 | 99.1 |
| Immundiagnostik/Roche | 226 | 2/224 | 99.1 |
| Immundiagnostik/Abbott | 226 | 2/224 | 99.1 |
